# The association between mental status, personality traits, and discrepancy in social isolation and perceived loneliness among community dwellers

**DOI:** 10.1101/2022.06.29.22277075

**Authors:** Kumi Watanabe-Miura, Takuya Sekiguchi, Mihoko Otake-Matsuura

## Abstract

**Objectives:** To examine the factors associated with social asymmetry, which refers to the discrepancy between actual social isolation and perceived loneliness, focusing on an individual’s mental status and personality traits.

**Methods:** This study introduced a cross-sectional study design that was utilizing the data from the University of Michigan Health and Retirement Study (HRS) from waves during 2014 and 2016. The participants were community dwellers aged 50 years and older. The outcome measurement, social asymmetry, was defined as the discrepancy between social isolation according to six criteria and loneliness as assessed by the three-item version of the Revised UCLA Loneliness Scale. Multinomial logistic regression models were conducted to examine the factors associated with social asymmetry.

**Results:** Fewer depressive symptoms and higher extraversion were consistently associated with social asymmetry, compared with both isolation and loneliness. Participants with higher neuroticism were likely to be lonely even in the absence of isolation, whereas those with lower neuroticism were likely to not be lonely even with social isolation. In addition, participants with fewer depressive symptoms, lower neuroticism, and higher extraversion were more likely to be not lonely even with social isolation, compared with lonely even in the absence of isolation.

**Conclusions:** Mental status and personality traits may closely relate to social asymmetry. This study suggests that incorporating social, mental, and psychological factors may be essential for interventions in social isolation and loneliness.

**Highlights:** Social asymmetry is a phenomenon reflecting the discrepancy between actual social isolation and perceived loneliness. This gap between situation and emotion may be associated with health risks. However, little information is available on social asymmetry, and its related factors remain poorly understood. Thus, we examined the factors associated with social asymmetry using data from the University of Michigan Health and Retirement Study (HRS). We hypothesized that both mental status and personality traits play a role in social asymmetry due to their effect on individuals’ perceptions.

As a result, depressive symptoms, neuroticism, and extraversion, as well as demographic and socioeconomic status were consistently associated with both social asymmetry outcomes: 1) only social isolation (without loneliness) and 2) only loneliness (without social isolation). This result implies that mental and psychological factors were associated with social asymmetry in a complex manner and that incorporating social, mental, and psychotherapeutic aspects in social interventions may be essential for future intervention strategies for social isolation and loneliness.

## 1. Introduction

Social isolation is a serious public concern. A National Health and Aging Trends Study found that 7.7 million older adults living in the community (24%) were socially isolated in the United States.^1^ Although the number of people with social isolation or loneliness is further increasing during the COVID-19 pandemic,^2-6^ there is a great deal of concern about the physical and mental health implications of social isolation and loneliness.^2,7-10^

Numerous empirical studies have revealed that social isolation and loneliness affect our health. Previous studies have reported the link to several health outcomes, such as depression and anxiety,^11-13^ mortality,^14-15^ and the onset of dementia.^16-17^ These studies are based on a variety of definitions and measurements of social isolation and loneliness. Social isolation often refers to objective isolation, which is a limited social relationship in structural (social networks) or functional (social support) facets.^14^ On the other hand, loneliness is generally assessed as subjective isolation that refers to situations in which an individual feels uncomfortable or has an unacceptable lack of social connectivity.^18^ Although social isolation and loneliness can co-occur, these are different concepts. Some people may not feel loneliness even during social isolation or may feel loneliness in the absence of social isolation. These discrepancies between social isolation and loneliness have been termed social asymmetry. ^19^

There is a paucity of studies on social asymmetry. Victor et al. classified social groups into four types of combination of social isolation and loneliness using data of the 1950s–1960s .^20^ However, the characteristics and factors associated with the classification of discrepancy have not been discussed. Other prior studies have reported relationships between the social asymmetry types and health outcomes. A study using the Irish Longitudinal Study on Ageing (TILDA) and the English Longitudinal Study of Aging (ELSA) categorized social asymmetry into “Concordantly Lonely & Isolated,” “Discordant: Robust to Loneliness,” and “Discordant: Susceptible to Loneliness.”^19^ The “Discordant: Robust to Loneliness” group that felt less lonely in the face of social isolation outperformed the “Discordant: Susceptible to Loneliness” group in all cognitive function tests. Moreover, a study from the ELSA classified six types based on living arrangement, loneliness, and social isolation using cluster analysis.^21^ Groups with either social isolation or loneliness were associated with health outcomes, including activities of daily living, subjective health status, chronic illness, and mental health. Another study using data from the Canadian Longitudinal Study of Aging clarified the four categories of combinations of social isolation and loneliness and found relationships with social support and psychological stress.^22^

To summarize, some prior studies have suggested that the social asymmetry types, as well as social isolation and loneliness alone, may be key factors for various health risks. Although there is still limited literature on social asymmetry, more evidence is required to be accumulated for the development of the intervention in social isolation and loneliness. Specifically, understanding the factors associated with social asymmetry has clinical usefulness because these factors possibly buffer the effects of interventions or may be effective in intervening themselves on social isolation and loneliness. We hypothesized that mental status and personality traits are associated with social asymmetry due to their effect on individuals’ perceptions, which create gaps in their actual situation of social isolation and perception of loneliness. While little is available on social asymmetry, especially the role of personality traits, our study may provide new empirical knowledge and is expected to offer new insight into mental health and social activity interventions.

Therefore, this study aimed to examine the comprehensive factors associated with social asymmetry, especially focusing on mental status and personality traits.

## 2. Materials and methods

### 2.1 Study design and setting

This study introduced a cross-sectional study design that utilized data from the University of Michigan Health and Retirement Study (HRS), which is a biennial longitudinal study with a nationally representative sample. The HRS includes noninstitutionalized adults over the age of 50 and their spouses selected via multistage probability sampling.

We analyzed the RAND HRS Longitudinal File^23^ and linked RAND files with HRS Leave Behind Questionnaires (LBQ)^24^ which include our interest variables, such as loneliness and social isolation. The LBQ introduced a rotating random subsample of the longitudinal panel. A random subsample of 50% of the HRS participants (Subsample A) responded in the relevant year, and the other 50% (Subsample B) responded in the next survey. Thus, we combined the 2014 (Subsample A) and 2016 (Subsample B) waves to obtain the whole sample. Information about e ducational attainment and institutionalized status was obtained from the Cross-Wave Tracker File.

The participants aged 50 and older, noninstitutionalized adults, and those who answered the items related to demographics (age, sex, educational attainment, and race) were included in the analysis. Participants who were missing all indicators on the LBQ, including loneliness, social isolation indicators, and personality traits, were excluded. First, 18,899 participants who were aged over 50 and answered the LBQ (who were provided LBQ weight) during the 2014 or 2016 waves were initially enrolled. After participants were excluded due to institutionalization, we had 17,948 total participants, and ultimately, 13,094 participants with no missing values regarding demographics were included in the final analysis.

### 2.2 Measurement

#### 2.2.1 Social isolation

Although a variety of definitions and measurements for social isolation exist with no golden standard, we used criteria with reference to previous research in the ELSA and the HRS. ^25-28^ We assigned one point for each of six criteria: (i) lived alone, (ii) unmarried, (iii)-(v) had less than monthly contact with family, children, and friend, and (vi) had less than monthly participation in any social groups or organizations. The score ranged from 0–6, and a higher rating corresponded to severe isolation. Given the prevalence of social isolation,^1^ the top quartile of the scores (score ≥3) was treated as socially isolated.

#### 2.2.2 Loneliness

Loneliness was measured using a three-item version of the Revised UCLA Loneliness Scale.^29^ This scale assesses the frequency of feeling loneliness: (i) lacking companionship, (ii) feeling left out, and (iii) being isolated from others. Each question has three options, including “never or hardly ever,” “some of the time,” or “often,” with 1—3 points given for these responses. The total score ranged from 3—9 by summing the scores for each item. A high score corresponds to severe loneliness. Considering the prevalence of loneliness,^30^ the top quartile of the scores (score ≥6) was treated as loneliness.

#### 2.2.3 Social asymmetry

The social asymmetry, which is our main interest, were represented by a matrix between social isolation and loneliness. Based on the categorization of social isolation and loneliness divided into quintiles as described earlier, we classified the combination of social isolation and loneliness into four categories: (i) No social isolation and loneliness (No SL), (ii) Only social isolation (Only S), (iii) Only loneliness (Only L), and (vi) Social isolation and loneliness (SL). In those groups, we defined “Only S” and “Only L,” which show the discrepancy between social isolation and loneliness, as social asymmetry

### 2.3 Independent variables

#### 2.3.1 Demographics

Age was treated as a continuous variable. Sex was categorized as female and male. Education was dichotomized into high school or less and college or above. Race was divided into White/Caucasian, Black or African American, and other. Household income, which is the sum of the income of respondents and their spouses or partners, was treated as continuous ($1000/unit). Medical conditions were divided into yes (has) or no (does not have) for each disease status: diabetes, stroke, cancer (excluding skin cancer), and heart disease. IADL was measured by five items assessing any difficulty with preparing a meal, making phone calls, taking medications, paying the bills and keeping track of expenses, and shopping. Each question has several response options, including “yes,” “no,” “can’t do,” and “don’t do.” Participants who reported difficulties with at least one item (i.e., answered “yes” or “can’t do”) were defined as dependent.

#### 2.3.2 Mental and psychological status

Mental health status was measured by the eight-item short version of the Center for Epidemiologic Studies-Depression Scale (CES-D-8).^31^ This scale consists of eight items that assess experiencing depressive symptoms for much of the time during the past week (e.g., felt depressed, felt everything was an effort, and felt sleep was restless). Each item was recorded as yes or no and the total score ranged from 0–8, with high scores indicating severe depressive symptoms. The traditional broadly used cut-off for the CES-D 8 was a score of ≧3. However, previous work has suggested using a more conservative cut-off for the CES-D in the older population,^32^ and data in the HRS showed that a cut-off of ≥4 for the CES-D-8 may be equivalent to a cut-off of 16 for the original CES-D (20-item).^33^ Thus, we introduced a cut-off of ≥4 and dichotomized scores into cases of depression syndrome (≧ 4) or no depression syndrome. Neuroticism and extroversion from among the Big 5 Personality traits were measured by the items of the Midlife Development Inventory Personality Scales.^34^ Considering the multicollinearity that shows strong correlations with medium or large correlation coefficients among personality domains in these data, this study introduced the neuroticism and extraversion domains that were related to social isolation and loneliness in a previous study^35^ as personality factors. Neuroticism is the personality trait that describes negative affect, such as nervousness, moodiness, and temperament.^36^ Extraversion is a personality trait that describes active people with sociable, talkative, and assertive natures.^37^

### 2.4 Analysis

This study examined the comprehensive factors associated with social asymmetry, with a particular focus on mental status and personality traits.

First, participants were assigned to one of the four groups based on the classification of social isolation and loneliness. Then, a multinomial logistic regression model was utilized to determine the factors related to the social asymmetry groups; “Only S” (do not feel lonely even in social isolation) and “Only L” (feel lonely in the absence of social isolation) compared with “SL” (social isolation and loneliness). Subsequently, we examined the differences between “Only S” and “Only L” in the model with “Only L” set as the reference.

As a supplementary analysis, we applied a multinomial logistic regression model utilizing a top-10%tile cut-off threshold for social isolation and loneliness in relation to social asymmetry, meaning a greater focus on severe social isolation and loneliness.

Missing data in the multinomial logistic regression model were imputed by multiple imputations with the fully conditional specification method. We created five imputed datasets and integrated each result of the analysis.

The level of significance was set at 0.05. All statistical tests were performed using SAS version 9.4.

## Results

Table 1 shows the participants’ demographic characteristics by the social isolation and loneliness categories. The results showed that the groups with social isolation (“Only S” and “SL”) involved older participants compared with those without isolation. The groups with social isolation, loneliness, or both had higher proportions of females than the “No SL” group. The “SL” group had the highest prevalence of depressive symptoms and IADL dependence.

**Table 1.**
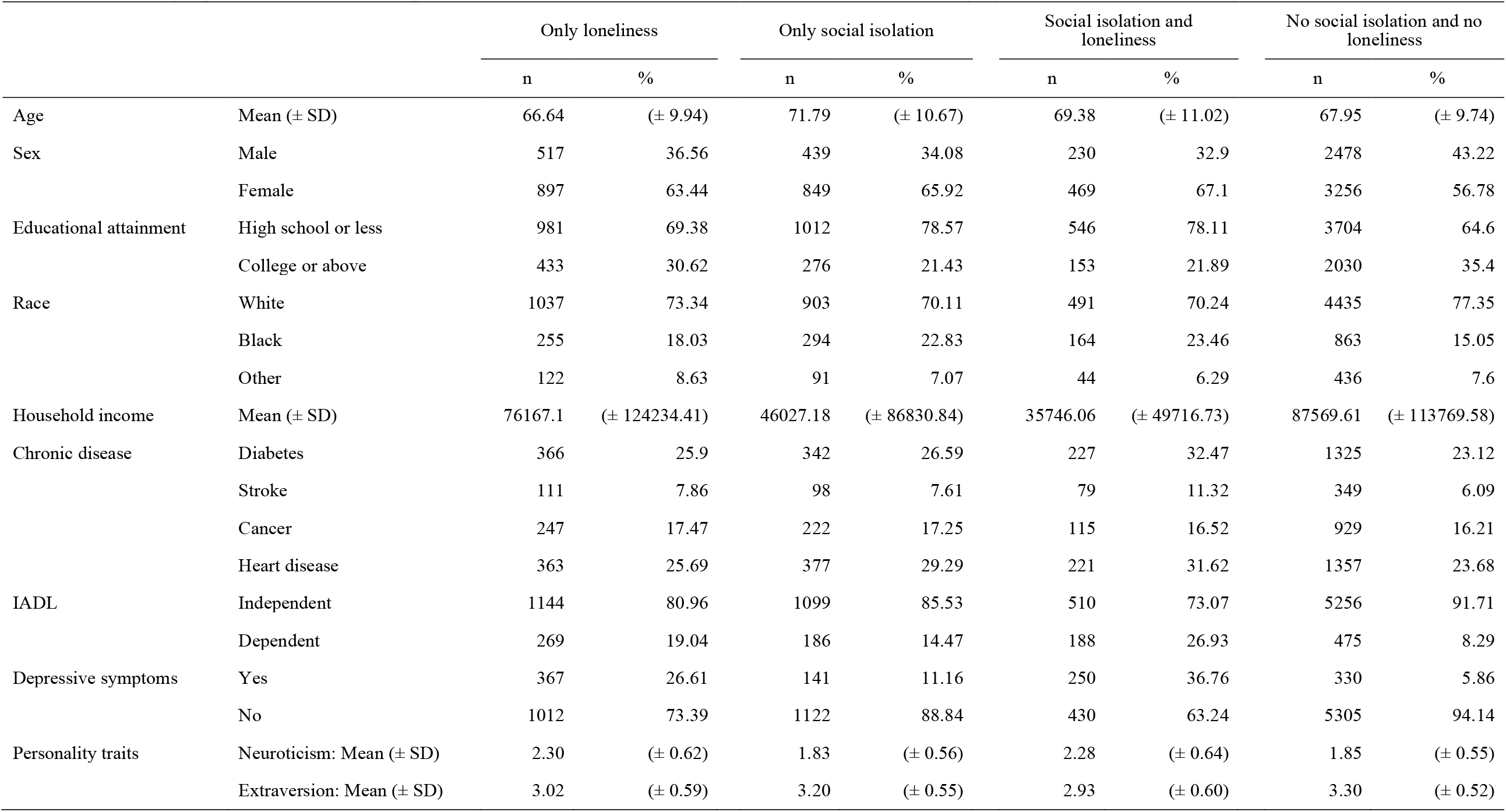
Demographic statistics of independent variables by social isolation and loneliness categories

Table 2 describes the results of multinomial logistic regression analysis for the combination of social isolation and loneliness, with the “SL” group set as the reference. Depressive symptoms and personality traits were closely associated with all three groups. Participants with a less depressive tendency and higher extraversion were more likely to be in social asymmetry groups than “SL.”. Neuroticism was associated with social asymmetry as well; however, the direction of the association differed. While higher neuroticism was likely to be “Only L,” lower neuroticism was likely to be “Only S.”

**Table 2.** Multinomial logistic regression analysis for the combination of social isolation and loneliness with SL set as the reference

|  |  | OR | 95% CI |  |  |
| --- | --- | --- | --- | --- | --- |
| No isolation and loneliness (No SL) | Age (Continuous) | 0.990* | 0.981 | - | 0.998 |
|  | Sex (ref. Male) | 0.804** | 0.691 | - | 0.937 |
|  | Educational attainment (ref. College or above) | 1.127 | 0.929 | - | 1.369 |
|  | Race: Black (ref. White) | 0.621*** | 0.503 | - | 0.767 |
|  | Race: Other (ref. White) | 1.325 | 0.990 | - | 1.772 |
|  | Household income (Continuous) | 1.010*** | 1.007 | - | 1.012 |
|  | Depressive symptoms (ref. No depression) | 0.215*** | 0.180 | - | 0.256 |
|  | IADL (ref. Independent) | 0.708*** | 0.594 | - | 0.844 |
|  | Diabetes (ref. No) | 0.944 | 0.804 | - | 1.108 |
|  | Stroke (ref. No) | 0.788 | 0.598 | - | 1.037 |
|  | Cancer (ref. No) | 1.208 | 0.990 | - | 1.472 |
|  | Heart disease (ref. No) | 0.983 | 0.828 | - | 1.166 |
|  | Neuroticism (Continuous) | 0.463*** | 0.407 | - | 0.528 |
|  | Extraversion (Continuous) | 2.227*** | 1.881 | - | 2.635 |
| Only social isolation (Only S) | Age (Continuous) | 1.021*** | 1.010 | - | 1.032 |
|  | Sex (ref. Male) | 1.113 | 0.928 | - | 1.333 |
|  | Educational attainment (ref. College or above) | 1.469*** | 1.186 | - | 1.819 |
|  | Race: Black (ref. White) | 0.999 | 0.790 | - | 1.263 |
|  | Race: Other (ref. White) | 1.512* | 1.028 | - | 2.225 |
|  | Household income (Continuous) | 1.004*** | 1.002 | - | 1.007 |
|  | Depressive symptoms (ref. No depression) | 0.381*** | 0.305 | - | 0.476 |
|  | IADL (ref. Independent) | 0.912 | 0.721 | - | 1.153 |
|  | Diabetes (ref. No) | 0.956 | 0.781 | - | 1.170 |
|  | Stroke (ref. No) | 0.756 | 0.548 | - | 1.043 |
|  | Cancer (ref. No) | 1.090 | 0.847 | - | 1.402 |
|  | Heart disease (ref. No) | 1.032 | 0.835 | - | 1.275 |
|  | Neuroticism (Continuous) | 0.398*** | 0.341 | - | 0.463 |
|  | Extraversion (Continuous) | 1.571*** | 1.299 | - | 1.901 |
| Only loneliness (Only L) | Age (Continuous) | 0.982** | 0.971 | - | 0.992 |
|  | Sex (ref. Male) | 0.913 | 0.767 | - | 1.088 |
|  | Educational attainment (ref. College or above) | 1.102 | 0.884 | - | 1.375 |
|  | Race: Black (ref. White) | 0.867 | 0.662 | - | 1.136 |
|  | Race: Other (ref. White) | 1.257 | 0.899 | - | 1.758 |
|  | Household income (Continuous) | 1.010*** | 1.007 | - | 1.012 |
|  | Depressive symptoms (ref. No depression) | 0.564*** | 0.467 | - | 0.682 |
|  | IADL (ref. Independent) | 0.832 | 0.683 | - | 1.013 |
|  | Diabetes (ref. No) | 0.948 | 0.787 | - | 1.141 |
|  | Stroke (ref. No) | 0.826 | 0.582 | - | 1.173 |
|  | Cancer (ref. No) | 1.273* | 1.017 | - | 1.594 |
|  | Heart disease (ref. No) | 0.982 | 0.798 | - | 1.207 |
|  | Neuroticism (Continuous) | 1.192* | 1.021 | - | 1.391 |
|  | Extraversion (Continuous) | 1.222* | 1.012 | - | 1.477 |

Ref: Social isolation and Loneliness (SL)
\*P < 0.05; \*\* P< 0.01; \*\*\* P < 0.001
OR: odds ratio; IC: confidence interval

Demographic factors, including age and higher household income, were also consistently related to social asymmetry. While being younger was more likely to be “Only L,” being older was more likely to be “Only S” than “SL.” In addition, lower educational attainment had a higher likelihood of being “Only S,” and history of cancer had a higher likelihood of being “Only L.”

Table 3 describes the results of multinomial logistic regression analysis with the “Only L” group set as the reference to determine the differences in characteristics among the “Only S” and “Only L” groups. Participants with being older, being female, lower educational attainment, lower income, lower depressive symptoms, lower neuroticism, and higher extraversion were more likely to be “Only S” than “Only L.”

**Table 3.** Multinomial logistic regression analysis for the combination of social isolation and loneliness with Only L set as the reference

|  |  | OR | 95% CI |  |  |
| --- | --- | --- | --- | --- | --- |
| No isolation and loneliness (No SL) | Age (Continuous) | 1.008** | 1.002 | - | 1.014 |
|  | Sex (ref. Male) | 0.881* | 0.786 | - | 0.988 |
|  | Educational attainment (ref. College or above) | 1.023 | 0.908 | - | 1.152 |
|  | Race: Black (ref. White) | 0.717*** | 0.613 | - | 0.838 |
|  | Race: Other (ref. White) | 1.053 | 0.860 | - | 1.290 |
|  | Household income (Continuous) | 1.000 | 1.000 | - | 1.001 |
|  | Depressive symptoms (ref. No depression) | 0.381*** | 0.326 | - | 0.445 |
|  | IADL (ref. Independent) | 0.851* | 0.727 | - | 0.996 |
|  | Diabetes (ref. No) | 0.996 | 0.872 | - | 1.138 |
|  | Stroke (ref. No) | 0.953 | 0.762 | - | 1.192 |
|  | Cancer (ref. No) | 0.948 | 0.817 | - | 1.101 |
|  | Heart disease (ref. No) | 1.001 | 0.878 | - | 1.141 |
|  | Neuroticism (Continuous) | 0.389*** | 0.353 | - | 0.428 |
|  | Extraversion (Continuous) | 1.821*** | 1.641 | - | 2.022 |
| Only social isolation (Only S) | Age (Continuous) | 1.040*** | 1.031 | - | 1.049 |
|  | Sex (ref. Male) | 1.218* | 1.045 | - | 1.420 |
|  | Educational attainment (ref. College or above) | 1.332** | 1.118 | - | 1.588 |
|  | Race: Black (ref. White) | 1.152 | 0.939 | - | 1.412 |
|  | Race: Other (ref. White) | 1.203 | 0.912 | - | 1.587 |
|  | Household income (Continuous) | 0.995*** | 0.993 | - | 0.996 |
|  | Depressive symptoms (ref. No depression) | 0.675*** | 0.548 | - | 0.830 |
|  | IADL (ref. Independent) | 1.096 | 0.884 | - | 1.359 |
|  | Diabetes (ref. No) | 1.009 | 0.855 | - | 1.190 |
|  | Stroke (ref. No) | 0.915 | 0.667 | - | 1.256 |
|  | Cancer (ref. No) | 0.856 | 0.707 | - | 1.037 |
|  | Heart disease (ref. No) | 1.051 | 0.898 | - | 1.230 |
|  | Neuroticism (Continuous) | 0.334*** | 0.293 | - | 0.380 |
|  | Extraversion (Continuous) | 1.286*** | 1.126 | - | 1.468 |
| Social isolation and Loneliness (SL) | Age (Continuous) | 1.019** | 1.008 | - | 1.029 |
|  | Sex (ref. Male) | 1.095 | 0.919 | - | 1.304 |
|  | Educational attainment (ref. College or above) | 0.907 | 0.728 | - | 1.131 |
|  | Race: Black (ref. White) | 1.153 | 0.880 | - | 1.511 |
|  | Race: Other (ref. White) | 0.795 | 0.569 | - | 1.112 |
|  | Household income (Continuous) | 0.991*** | 0.988 | - | 0.993 |
|  | Depressive symptoms (ref. No depression) | 1.772*** | 1.467 | - | 2.139 |
|  | IADL (ref. Independent) | 1.202 | 0.987 | - | 1.463 |
|  | Diabetes (ref. No) | 1.055 | 0.876 | - | 1.271 |
|  | Stroke (ref. No) | 1.210 | 0.853 | - | 1.717 |
|  | Cancer (ref. No) | 0.785* | 0.627 | - | 0.983 |
|  | Heart disease (ref. No) | 1.019 | 0.829 | - | 1.253 |
|  | Neuroticism (Continuous) | 0.839* | 0.719 | - | 0.979 |
|  | Extraversion (Continuous) | 0.818* | 0.677 | - | 0.988 |

Ref: Only loneliness (Only L)
\*P < 0.05; \*\* P< 0.01; \*\*\* P < 0.001
OR: odds ratio; IC: confidence interval

Supplementary Tables 1 and 2 describe the models, focusing on severe social isolation and loneliness utilizing top 10%tile cut off. In the model with the “SL” group set as the reference (Supplementary Table 1), the significance of depressive tendency and neuroticism remained for the “Only S” group, consistent with Table 2; however, an association between personality traits and “Only L” was not observed. In the model with the “Only L” group set as the reference (Supplementary Table 2), participants with lower depressive tendency and lower neuroticism were still likely to be “Only S” than “Only L” while extraversion was not associated with.

## 4. Discussion

Although social isolation and loneliness are important and urgent public health issues, the factors associated with social asymmetry, which refers to the gap between the actual situation of social isolation and feeling lonely, have remained unknown. To the best of our knowledge, the current study is the first to examine comprehensive factors related to social asymmetry with a focus on the roles of mental status and personality traits using large-scale data. While previous studies have reported the factors associated with social isolation and loneliness alone ^1, 38-40^ our findings may add new knowledge and insight into social isolation and loneliness prevention by directing a spotlight on social symmetry as follows:

First, depressive symptoms and personality traits were consistently associated with both social asymmetry groups. While social isolation may closely be related to situational loneliness caused by environmental factors, internal loneliness—the perception of being alone that makes a person lonely—is influenced by psychological factors such as mental distress and personality traits.^41^ Although the association between mental health and social isolation and loneliness is well established, ^11-13, 42^ our work added that mental health is associated with social asymmetry, as are loneliness and social isolation individually. Further, the result showed that the personality traits, including neuroticism and extraversion, were related to social asymmetry.

To summarize the results regarding personality traits, higher extraversion was associated with two social asymmetry groups: “Only S” and “Only L.” Given that the extraversion is a personality trait that elicits individuals’ activeness in terms of a sociable, talkative, and assertive nature,^37^ the groups with either social isolation or loneliness may tend to be more extroverted than the combination social isolation–loneliness group.

Interestingly, the direction of the association between neuroticism and the two social asymmetry groups differed; higher neuroticism was associated with “Only L,” and lower neuroticism was associated with “Only S.” Furthermore, our model with set “Only L” as reference suggested that “Only S” had significantly higher extraversion compared with “Only L” even though “Only S” has limited social interaction.

Neuroticism and extraversion are personality traits that may be closely associated with stress reactivity and resilience in daily life. A prior study suggested that people with higher neuroticism may experience more event-related distress and have greater stress-reactivity than those with lower neuroticism.^43^ Another study reported that higher neuroticism and lower extraversion were related to worse adaptation to the COVID-19 lockdown,^44^ which is a situation similar to social isolation. Considering those reports, our result implies that the “Only S” group, which does not feel lonely in an isolated situation, may feel less loneliness because of lower stress-reactivity via lower neuroticism, while for the “Only L” group, the situation is reversed. In addition, people with higher extraversion may be resistant to stressful situations.^45^ Significantly higher extraversion in the “Only S” group compared with the “Only L” group might help these individuals cope with loneliness by facilitating their resistance to stressful situations, such as social isolation in this case. In summary, one possible hypothesis is that personality may be associated with one`s perception of isolation via resistance and stress reactivity and may sometimes create a discrepancy between the actual situation and perception of isolation.

It should also be noted that our supplementary analysis focusing on severe social isolation and loneliness showed that personality traits were not significantly associated with “Only L,” which is the group with severe loneliness despite the absence of severe isolation. Rather than personality traits, gender, household income, depressive symptoms, and cancer history were associated with “Only L,” although neuroticism was consistently associated with “Only S.” This result implies that socio-demographic factors, physical and mental health status rather than personality traits may be closely related to “Only L” in the case of severe loneliness without severe social isolation.

In addition to mental status and personality traits, demographic and socio-economic factors, including age, educational attainment, household income, and history of disease were also associated with social asymmetry. In those results, “Only S” which is without loneliness tended to be older and had lower educational attainment compared with “SL” which is a combination of high risk of social isolation and loneliness. This result seems to be in the opposite direction of the association with the prior studies which reported that being older and having lower educational attainment were risk factors for loneliness alone.^46^ However, the work by Schoenmakers et al, which examined the ways of coping in various situations where loneliness occurs, suggested that the regulative coping for loneliness —a coping path by lowering expectations about their relationships— may be more often in people who are older and with low educational attainment.^47^ Given this report, our result can be explained as follows: people who are older and have lower educational attainment might adjust their aspirations even during social isolation, whereas other people tend to try to remove the stressor by increasing social contacts. Future studies are needed to accumulate evidence on the direction of the association between demographic and socio-economic factors, including age and educational attainment, and social asymmetry.

From the perspective of social intervention, this study has strong implications for isolation and loneliness prevention. In recent years, much attention has been focused on interventions for social isolation and loneliness. According to comprehensive reviews,^48-50^ many trials have emphasized social contacts, and most of them have intervened in a single way (e.g., increase social contact, social skill training). Considering our results, social involvement intervention may help improve individuals’ social isolation and situational loneliness; however, intervention in a single way could not address social asymmetry, which was associated with mental status and personality traits in a complex manner. Thus, this study implies that incorporating social, mental, and psychotherapeutic aspects in social interventions, such as a combination of social involvement and cognitive-behavioral interventions, may be essential for future intervention strategies.

The strength of the current study included examining the factors related to social asymmetry using large-scale data with comprehensive variables including mental health and personality traits, as the strengths of the HRS implied introducing various indicators, including socio-economic, physical, mental, and psychological factors. The limitations of this study should also be noted. First, our results could not address the causal relationship between variables and longitudinal changes. The factors related to social asymmetry in this study, such as depressive symptoms, can be bidirectionally linked to social isolation and loneliness. Second, the results were based on self-administered questionnaires. Self-reported bias, such as feelings of loneliness being influenced by the emotions at the time of completing the questionnaire, cannot be ruled out. Third, differences in evaluation between the clinical evaluation and self-reported indicators are possible in terms of the mental and psychological factors, including depressive symptoms, loneliness, and personality traits. Based on these limitations, future studies are expected to accumulate more evidence on social asymmetry.

## 5. Conclusions

The current study examined the association between mental status, personality traits, and social asymmetry among community-dwellers using the HRS dataset. As we hypothesized, depressive symptoms and personality traits were consistently associated with social asymmetry. The results indicated the possibility that mental status and personality traits are closely involved in creating gaps between social isolation and perceived loneliness. Our findings provided implications for social interventions, indicating that social, mental, and psychotherapeutic aspects may be essential for intervention. As this study has its set of limitations, future studies are expected to delve deeper into the mechanism of social asymmetry.

## Data Availability

All data produced are available at the Health and Retirement Study HP.

---

**Supplement 1.**
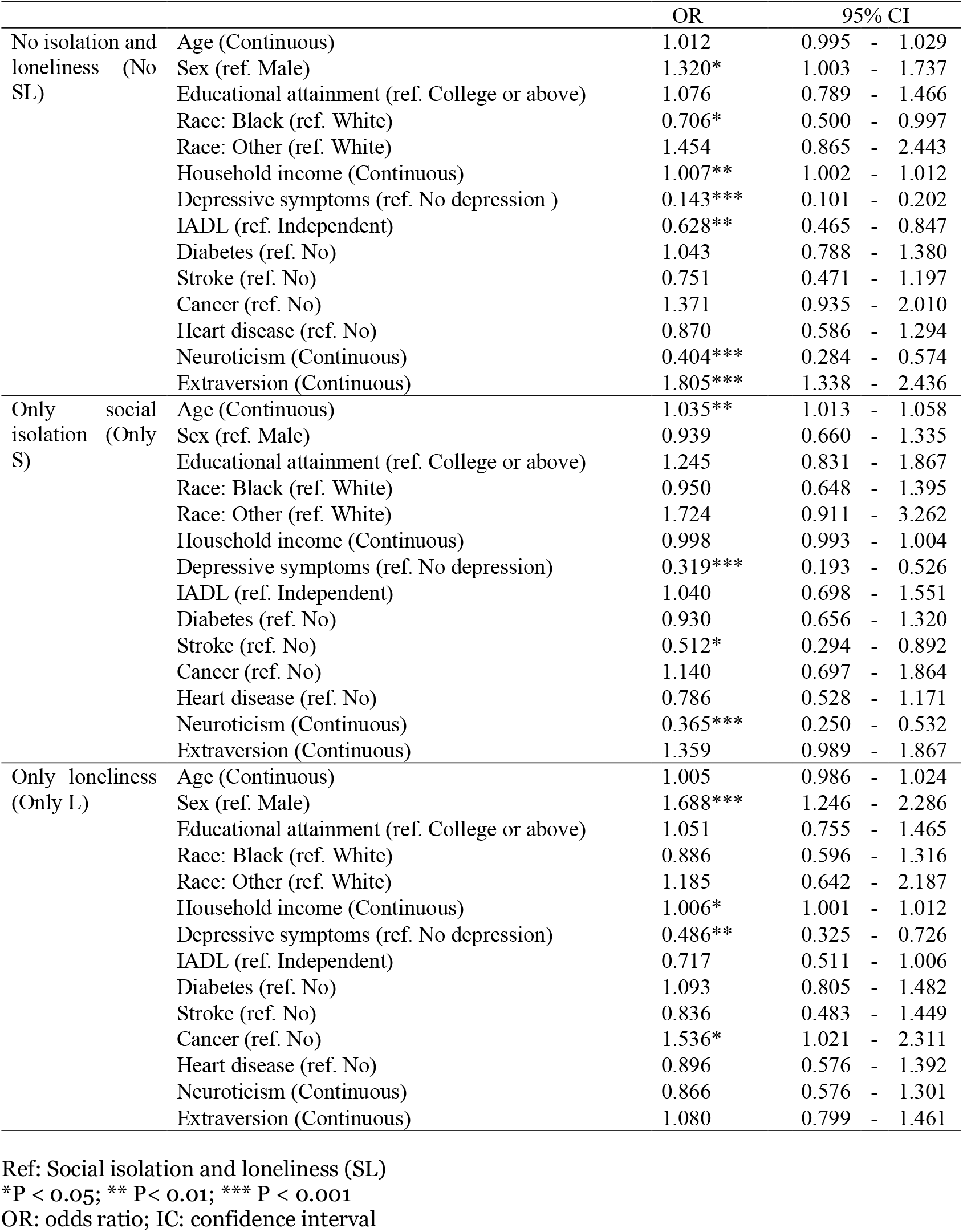
Multinomial logistic regression analysis for the combination of severe social isolation and loneliness with SL set as the reference

**Supplement 2.**
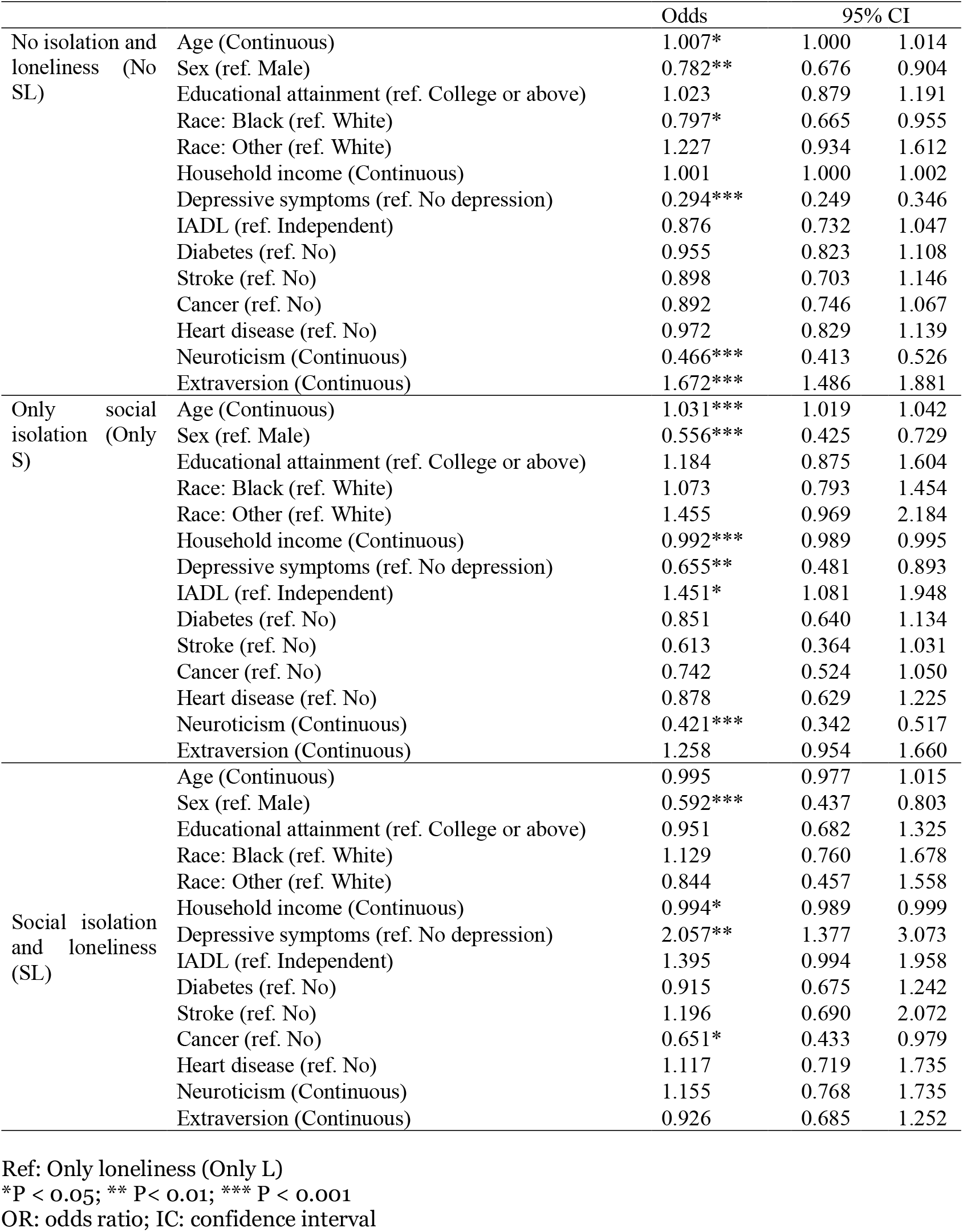
Multinomial logistic regression analysis for the combination of severe social isolation and loneliness with Only L set as the reference

